# The effect of neoadjuvant therapy on PD-L1 expression and CD8 lymphocyte density in non-small cell lung cancer

**DOI:** 10.1101/2022.04.11.22273684

**Authors:** Philipp Zens, Corina Bello, Amina Scherz, Michael von Gunten, Adrian Ochsenbein, Ralph A Schmid, Sabina Berezowska

**Author notes:** **Corresponding Author** Prof. Dr. Sabina Berezowska, Department of Laboratory Medicine and Pathology, Institute of Pathology, Lausanne University Hospital and Lausanne University, Rue du Bugnon 25, 1011 Lausanne, Switzerland. Department for anesthesiology, Hospital Grabs, Spitalstrasse 44, CH-9472 Grabs, Switzerland. Preliminary results have been presented at the World Conference on Lung Cancer, September 7th – 10th 2019 in Barcelona, Spain.

## Abstract

PD-L1 expression is the routine clinical biomarker for the selection of patients to receive immunotherapy in non-small cell lung cancer (NSCLC). However, the application and best timing of immunotherapy in the resectable setting is still under investigation. We aimed to study the effect of chemotherapy on PD-L1 expression and tumor infiltrating lymphocytes (TIL), which is to date still poorly understood. This retrospective, single-centre study cohort comprised 96 consecutive patients with NSCLC resected in 2000-2016 after neoadjuvant therapy, including paired chemo-naïve specimens in 57 cases. A biologically matched control cohort of 114 primary resected cases was included. PD-L1 expression, CD8+ TIL density and tertiary lymphoid structures were assessed on whole slides and correlated with clinico-pathological characteristics and survival.

Seven/57 and 12/57 cases had lower respectively higher PD-L1 expressions after neoadjuvant therapy. Most cases (n = 38) had no changes in PD-L1 expression and the majority of these showed PD-L1 < 1% in both samples (23/38 [60.5%]). CD8+ TILs density was significantly higher after chemotherapy (p = 0.031) in paired resections. Neoadjuvant cases showed no difference in PD-L1 expression or CD8+ TILs density compared to the chemotherapy naïve control cohort. In univariable analyses, higher CD8+ TILs density, higher numbers of tertiary lymphoid structures but not PD-L1 expression were significantly associated with better survival. Increased PD-L1 expression after neoadjuvant chemotherapy was visually associated with worse 5-year survival, lacking statistical significance probably due to the low number of cases. PD-L1 expression is mostly unchanged after neoadjuvant chemotherapy. However, an increase of PD-L1 expression after neoadjuvant therapy could be associated with worse survival.

## Introduction

Lung cancer is the deadliest cancer worldwide, mainly explainable by the late diagnosis due to presentation in advanced stages (UICC/AJCC TNM stage III/IV) (1). For early-stage non-small cell lung cancer (NSCLC), surgery still offers the best chance of cure (2). However, especially in nodal-positive patients, there is a high risk of recurrence and death. Since randomized trials have shown that additional neoadjuvant or adjuvant chemotherapy leads to better outcomes compared to resection only, it is generally accepted that patients with lymph node metastatic NSCLC should not receive surgery as a stand-alone treatment (3, 4). Adjuvant platinum-based chemotherapy offers a modest 5-year survival benefit of about 5% and is recommended for patients with completely resected early-stage, high-risk NSCLC – weighing the benefits and risks (5). Perioperative therapeutic approaches are a hotly debated topic, with immunotherapy-based combinations and targeted treatments – in EGFR mutated NSCLC – dominating the current trial landscape. According to surgical outcomes from the phase III CheckMate 816 trial, the neoadjuvant combination of nivolumab and chemotherapy showed considerably lower rates of residual tumor compared with chemotherapy alone (6). Primary results of the phase III global IMpower010 trial demonstrated a significant improvement in disease-free survival (DFS) comparing atezolizumab with best supportive care after adjuvant chemotherapy for resected stage IB – IIIA NSCLC. The greatest benefit was observed in patients with a tumoral programmed cell death 1 ligand 1 (PD-L1) expression (TPS) ≥1% (7).

Programmed cell death protein 1 (PD1) and its ligand PD-L1 belong to the costimulatory pathway of the adapted immune system (8). Extensive studies have explained the hijacking of this regulatory pathway by different tumor entities including lung cancer (8). PD1 is expressed on the surface of T cells, mediating inhibitory and stimulatory signals (8). The overexpression of PD-L1 on the surface of tumor cells leads to evasion of an appropriate tumor-induced response of the immune system by T cell apoptosis and exhaustion (8). The combination of immune checkpoint blockade and chemotherapy seems to be beneficial especially in patients with low levels of PD-L1 expressing tumor cells and ongoing trials are reporting positive results of this regimen in patients with resectable lung cancer (6, 9). However, the selection of appropriate patients is currently based only on PD-L1 expression in tumor tissue prior to medication, which is a suboptimal biomarker. More selective tools or, conceivably, a combination of multiple tumor immunity markers such as tumor mutational burden or CD8+ tumor infiltrating lymphocytes (TILs) are needed to predict response to treatment. This is highlighted by recent examples of immune checkpoint blockade (ICB) benefit irrespective of tumoral PD-L1 expression (10). Furthermore, it is still unclear how classic neoadjuvant chemotherapy influences the tumor microenvironment and if it could promote therapeutic ICB. Regarding the neoadjuvant setting, several studies have reported dynamic changes, though without a clear trend of altered PD-L1 expression after chemotherapy (Table 1) (11-22). These results contrast preclinical data substantiating an immunogenic effect to some chemotherapeutic agents and functional studies reporting the mechanisms involved in chemotherapy resistance and PD-L1 upregulation (14, 19, 23).

**Table 1:** Currently published studies investigating the effect of neoadjuvant regimens on PD-L1 expression and the immune microenvironment.

| study | year | number of patients | histology | paired samples (pre-/post-neo-adjuvant) | neoadjuvant treatment | PD-L1 clone | PD-L1 evaluation | PD-L1 change | negative/no change | CD8 clone | CD8 evaluation | CD8 change |
| --- | --- | --- | --- | --- | --- | --- | --- | --- | --- | --- | --- | --- |
| Remark [11] | 2016 | 21 | LUSC/LUAD/LCC | yes | chemotherapy | E1L3N | Semi-quantitative | no change | - | SP16 | manually | no change |
| Sheng [12] | 2016 | 32 | LUSC/LUAD | yes | chemotherapy | E1L3N | Semi-quantitative | decrease | - | - | - | - |
| Song [13] | 2016 | 76 | LUSC | yes | chemotherapy | PD-L1 | Semi-quantitative | increase | 31/65 | - | - | - |
| Zhang [14] | 2016 | 30 | LUSC/LUAD | yes | chemotherapy | Abcam | Semi-quantitative | - | - | - | - | - |
| Fujimoto [15] | 2017 | 35 | LUSC/LUAD | yes | chemoradiotherapy | 28-8 | TPS | decrease | 11/15 | C8/144B | Semi-quantitative | increase |
| Parra [16] | 2018 | 112 | LUSC/LUAD | no | chemotherapy | E1L3N* | automated | increase | - | C8/144B (fluorescent) | automated | no change |
| Rojko [17] | 2018 | 41 | LUSC/LUAD/NSCLC/SCLC | yes | chemotherapy | SP142 | Semi-quantitative | no change | -/29 | H&E | Semi-quantitative | no change |
| Choe [18] | 2019 | 33 | LUSC/LUAD | yes | chemoradiotherapy | 22C3<br>SP263 | TPS | no change | 13/16 | C8/144B | Semi-quantitative | increase |
| Fournel [19] | 2019 | 39 | LUSC/LUAD/LCNEC | yes | chemotherapy | E1L3N | TPS | increase | -/8 | SP16 | automated | no change |
| Guo [20] | 2019 | 63 | LUSC/LUAD/LCNEC | yes | chemotherapy | 22C3 | TPS | increase | -/40 | - | - | - |
| Shin [21] | 2019 | 86 | LUSC/LUAD | yes | chemotherapy | E1L3N | TPS | increase | 15/40 | - | - | - |
| Yoneda<br>[22] | 2019 | 41 | LUSC/LUAD | yes | Chemotherapy,<br>chemoradiotherapy | E1L3N | TPS | increase<br>(CCRT),<br>no<br>change<br>(CT)<br>no<br>change<br>(CT) | - | C8/144B | manually | increase |

LUSC, lung squamous cell carcinoma; LUAD, lung adenocarcinoma; LCC, large cell carcinoma; NSCLC, non-small cell lung carcinoma; CCRT, concurrent chemoradiotherapy; CT, chemotherapy ; \*, fluorescent

Here, we aimed to assess PD-L1 expression and CD8+ TILs density and their prognostic importance in a real-life cohort of patients with NSCLC resected after neoadjuvant chemo(radio)therapy compared to paired diagnostic samples and a biologically matched control cohort with primary resected advanced NSCLC.

## Material and Methods

### Study population

This retrospective single center study was conducted on consecutive patients with NSCLCs, resected between January 2000 and December 2016 in the Department of thoracic surgery of the Inselspital and diagnosed at the Institute of Pathology, University of Bern as previously described (24). It includes a study cohort of cases resected after neoadjuvant therapy and a biologically matched control cohort of primary resected cases of lung squamous cell carcinoma (LUSC) and lung adenocarcinoma (LUAD) at a locally advanced stage, defined by the presence of mediastinal lymph-node metastases (pN2) (25). The cases were included according to pathology reports, validated and expanded by considering the clinical files of the Inselspital Bern (clinical data), cantonal cancer registry of Bern (survival data) and by contacting the general practitioners (clinical and survival data).

The initial study cohort consisted of 131 cases and was reduced to 119 cases corresponding to 118 patients after excluding tumors with neuroendocrine histology and patients not treated with neoadjuvant intention. Finally, 22/118 patients were excluded due to insufficient residual tumor in any of the diagnostic blocks resulting in 96 patients included for evaluation of tumoral PD-L1 expression and CD8+ TILs and 95 patients for evaluation of tertiary lymphoid structures (TLS). Regarding pre-neoadjuvant therapy specimens, 86/118 patients had available diagnostic biopsies or cytology specimens, in 57 cases with sufficient tumor content for PD-L1 assessment and in 36 cases with adequate material for CD8+ TILs evaluation, excluding cytologies and lymph node biopsies without desmoplastic reaction. TLS were not evaluated in the pretherapeutic specimens.

The initial control cohort consisted of 115 cases including 60 patients with LUAD and 55 patients with LUSC. One case was excluded from further evaluation because of insufficient tumor present in the diagnostic blocks. Finally, 114 patients were included for PD-L1 evaluation and 111 patients for CD8 and TLS evaluation (3 patients missing due to repeatedly insufficient scanning quality).

For harmonization, all cases were pathologically re-evaluated by SB and PZ and re-staged according to the current 8^th^ edition of the UICC TNM classification (25). Additionally, the predominant growth pattern was assessed for primary resected LUAD cases according to the current 2021 World Health Organization criteria (26). Table 2 summarizes the baseline characteristics of the cohorts.

**Table 2:** Baseline characteristics of the study and control cohort.

|  | Study Cohort<br>(n = 96) | Control Cohort<br>(n = 114) | p value |
| --- | --- | --- | --- |
| **age (median [IQR])** | 63.50 [55.75, 70.00] | 63.50 [58.00, 70.00] | **0.418°** |
| **sex (%)** | **n = 96 (%)** | **n = 114 (%)** | **0.553\*** |
| female | 28 (29.2) | 38 (33.3) |  |
| male | 68 (70.8) | 76 (66.7) |  |
| **smoking status (%)** | **n = 83 (%)** | **n = 90 (%)** | **0.429\*** |
| never/ex-smoker | 56 (67.5) | 55 (61.1) |  |
| active smoker | 27 (32.5) | 35 (38.9) |  |
| **histology (%)** | **n = 96 (%)** | **n = 114 (%)** | **0.678\*** |
| LUSC | 46 (47.9) | 54 (47.4) |  |
| LUAD | 47 (49.0) | 60 (52.6) |  |
| other | 3 (3.1) |  |  |
| **tumor size (median [IQR])** | 3.20 [2.00, 4.85] | 4.75 [3.00, 6.07] | **<0.001°** |
| **Major pathological response (%)** | **n = 96 (%)** |  |  |
| MPR | 33 (34.4) |  |  |
| No MPR | 63 (65.6) |  |  |
| **(y)pT (%)** | **n = 96 (%)** | **n = 114 (%)** | **0.027°** |
| (y)pT0 | 1 (1.0) |  |  |
| (y)pT1 | 32 (33.3) | 17 (14.9) |  |
| (y)pT2 | 22 (22.9) | 37 (32.5) |  |
| (y)pT3 | 20 (20.8) | 31 (27.2) |  |
| (y)pT4 | 21 (21.9) | 29 (25.4) |  |
| **stage (%)** | **n = 96 (%)** | **n = 114 (%)** |  |
| I | 17 (17.7) |  |  |
| II | 25 (26.0) |  |  |
| III | 50 (52.1) | 104 (91.2) |  |
| IV | 4 (4.2) | 10 (8.8) |  |
| **growth pattern (%)** |  | **n = 58 (%)** |  |
| acinar/papillary |  | 15 (25.9) |  |
| solid |  | 28 (48.3) |  |
| micropapillary |  | 15 (25.9) |  |
| **type of resection (%)** | **n = 96 (%)** | **n = 114 (%)** | **0.486\*** |
| wedge |  | 3 (2.6) |  |
| lobectomy | 53 (55.2) | 62 (54.4) |  |
| bilobectomy | 5 (5.2) | 8 (7.0) |  |
| pneumonectomy | 38 (39.6) | 41 (36.0) |  |
| **neoadjuvant therapy (%)** | **n = 94 (%)** |  |  |
| Cisplatin + Docetaxel | 54 (57.4) |  |  |
| Carboplatin + Paclitaxel | 5 (5.3) |  |  |
| Cisplatin + Pemetrexed | 13 (13.8) |  |  |
| Cisplatin + Gemcitabine | 8 (8.5) |  |  |
| Cisplatin + Vinorelbine | 5 (5.3) |  |  |
| Cisplatin + Etoposide | 1 (1.1) |  |  |
| Other | 8 (8.5) |  |  |
| **adjuvant therapy (%)** | **n = 88 (%)** | **n = 99 (%)** | **<0.001\*** |
| no | 65 (73.9) | 32 (32.3) |  |
| yes | 23 (26.1) | 67 (67.7) |  |

No statistical comparison of stage, growth patterns, residual tumor and neoadjuvant therapy due to inherent differences.
\* Fisher's exact test, ° Wilcoxon rank-sum test. Main variable names and p-values are bold. Statistically significant p-values are red.

This study was carried out according to the REMARK criteria and approved by the Cantonal Ethics Commission of the Canton of Bern (KEK 2017-00830), which waived the requirement for written informed consent (27).

### Survival Analyses

We restricted the survival analyses to five years after initial diagnosis to account for the multimorbidity of older patients. Overall survival (OS) was defined as the period from the beginning of treatment to death of any cause. DFS was defined as the period from the beginning of treatment to clinically reported relapse or death of any cause. The beginning of treatment was defined by the start of neoadjuvant therapy in the study cohort or the date of resection in the control cohort and in 2 cases with missing information about the starting date of neoadjuvant therapy. Patients with stage IV disease (n = 14), missing survival information (n = 7), non-curative resection (n = 2) or last follow-up information within 30 days after surgery (n = 12) were excluded from survival analyses resulting in 175 patients included (study cohort n = 83, control cohort n = 92). Median OS was 35 (95% CI 29 – NA) months and 87 events were observed (Supplementary figure 1A). Median DFS was 18 (95% CI 15 – 25) months and 118 events were observed (Supplementary figure 1B). There was no significant difference of survival between the study cohort and the control cohort (Supplementary figure 1C/1D).

### Immunohistochemical staining and scanning

For immunohistochemical staining appropriate tissue blocks were selected after screening all available H&E slides. PD-L1 staining was effectuated in a closed system using the Ventana SP263 assay (Roche Diagnostics International AG, Rotkreuz, Switzerland) on the fully automated immunostainer BenchMark ULTRA (Roche Diagnostics International AG) following the manufacturer’s instructions. The sections were pre-processed using CC1 buffer at 100°C for 64 minutes, followed by antibody incubation at 37°C for 16 minutes and visualization with DAB. CD8 staining was effectuated on a fully automated immunostainer BOND III (Leica Biosystems, Muttenz, Switzerland) using C8/144B (Agilent, Santa Clara, CA, United States). The sections were pre-processed with ER2 buffer at 100°C for 20 minutes, followed by incubation of the diluted antibody (1:200) for 15 minutes and visualization with DAB. Selected slides were digitized using the Pannoramic P250 Flash III (3DHistech, Budapest, Hungary) in multiple runs at a resolution of 0.2431 µm/pixel. The tissue had been obtained during the routine diagnostic workflow and the formalin-fixed paraffin-embedded tissue had been stored at the Institute of Pathology Bern according to the recommendation of the Swiss Society of Pathology (28). There was no evidence of time-dependent staining bias with similar distributions of PD-L1 or CD8 expression along the period of observation (Supplementary figure 2A/2B).

### PD-L1 assessment

Specimens with at least 100 tumor cells were eligible. PD-L1 expression was assessed by PZ and reviewed on a double-headed microscope together with SB. In cases of discordant assessment consensus was achieved. PD-L1 expression was assessed as the tumor proportional score (TPS), defined by the proportion of PD-L1 positive tumor cells of all tumor cells. PD-L1 positive tumor cells were defined as showing membranous staining of any intensity. TPS was assessed as a continuous parameter in 1% increments up to 10% and 5% increments in cases showing >10% expression. For statistical analyses, cases were assigned to the three clinically relevant bins of TPS <1%, 1 – 49% or ≥50%. PD-L1 positive cases were defined by TPS ≥1% and strong expressing cases were defined as TPS ≥50%.

### Assessment of CD8 positive tumor infiltrating lymphocytes and tertiary lymphoid structures

For the assessment of CD8+ TILs, only biopsies of non-lymph nodes or lymph nodes with desmoplastic reaction were eligible. We evaluated CD8+ TILs per mm^2^ applying a semi-automated approach using the open-source software QuPath (Supplementary figure 3) (29). First, we manually annotated regions of interest following recommendations of the International Immuno-Oncology Biomarkers Working Group (30, 31). Thus, only TILs within the borders of the invasive front of tumors were evaluated and smaller satellite nodules without desmoplastic reaction were not included in the assessment. In neoadjuvant cases with extensive fibrotic areas, only the stroma adjacent to the tumor nests was included for analyses. Next, cells in the annotated regions were segmented using the threshold-based watershed detection of QuPath followed by the application of a series of object classifiers for exclusion of anthracotic pigments and artefacts before classification and counting of CD8 negative and positive cells (technical manuscript in preparation). The performance of this automated detection and classification was compared in 22 cases using 5000 × 5000 px wide squares against manual counting of one observer (PZ, Supplementary table 1).

Regarding the evaluation of TLS, the digitized H&E sections were used for manual assessment of the number and activity (presence of germinal centers) of TLS in the resection specimens by PZ (32). In 44 cases, another block than used for PD-L1 or CD8 assessment was evaluated due to the presence of larger areas of adjacent normal lung tissue. All nodular aggregates of lymphocytes in the tumor region and within 7 mm of the tumor border were counted (32). In cases of densely infiltrated tumoral stroma, only nodular aggregates apparent on low magnification were included.

### Statistics

All analyses were conducted using R software (version 4.0.5, https://cran.r-project.org/) with suitable packages. For comparison of naturally ordered categorical variables or continuous variables, we used the Wilcoxon rank-sum or Kruskal-Wallis test and for comparison of other categorical variables the Fisher’s exact test. Correlation was assessed using the Spearman test. Survival analyses were conducted using the Log-rank test and univariable cox proportional hazard models. Kaplan-Meier plots were used for the representation of survival curves. Multivariable cox proportional hazard models were used for correction for confounders, which were selected based on a significance level of p ≤ 0.1. CD8 density was included as binary variable (low vs. high) in all survival models. It was dichotomized using maximally selected rank statistics based on Log-rank scores as test statistic and the approximation by Hothorn and Lausen for small sample sizes (33).

## Results

### No upregulation of PD-L1 expression by neoadjuvant therapy

After neoadjuvant therapy, PD-L1 expression was <1% in 43/96 (44.8%) cases, 1 - 49% in 31/96 (32.3%) cases and ≥50% in 22/96 (22.9%) cases. In the primary resected cohort, PD-L1 expression was <1% in 40/114 (35.1%) cases, 1 - 49% in 47/114 (41.2) cases and ≥50% in 27/114 (23.7%) cases. There was no significant difference in PD-L1 expression between the neoadjuvant cohort and the primary resected control cohort also after adjusting for histology.

Except for smoking status (active smoker vs. former-/ never smoker) none of the clinico-pathological parameters was associated with higher PD-L1 expression. In the neoadjuvant cohort, active smoking was associated with a higher PD-L1 TPS (p = 0.013). Active smokers in both cohorts had a significantly higher frequency of PD-L1 positive tumors (p_neoadjuvant_ = 0.02, p_control_ = 0.026, Supplementary table 2). PD-L1 expression was not significantly altered comparing paired pre-/post-neoadjuvant samples. Overall, 7/57 (12.3%) tumors had lower PD-L1 expression and 12/57 (21.1%) had higher PD-L1 expression in the resection specimen, as assessed regarding the clinically significant cut-offs of 1% and 50% (Figure 1, Supplementary table 3). Four/7 cases showed lowed PD-L1 expression regarding the 50% cut-off and 5/7 regarding the 1% cut-off (2 cases changed from ≥50% to <1%). A positive or negative change of PD-L1 could not be associated with response to neoadjuvant therapy (major pathological response [MPR] yes/no), patients’ sex, tumor histology or change of CD8 TILs density (Supplementary table 4).

**Figure 1:**
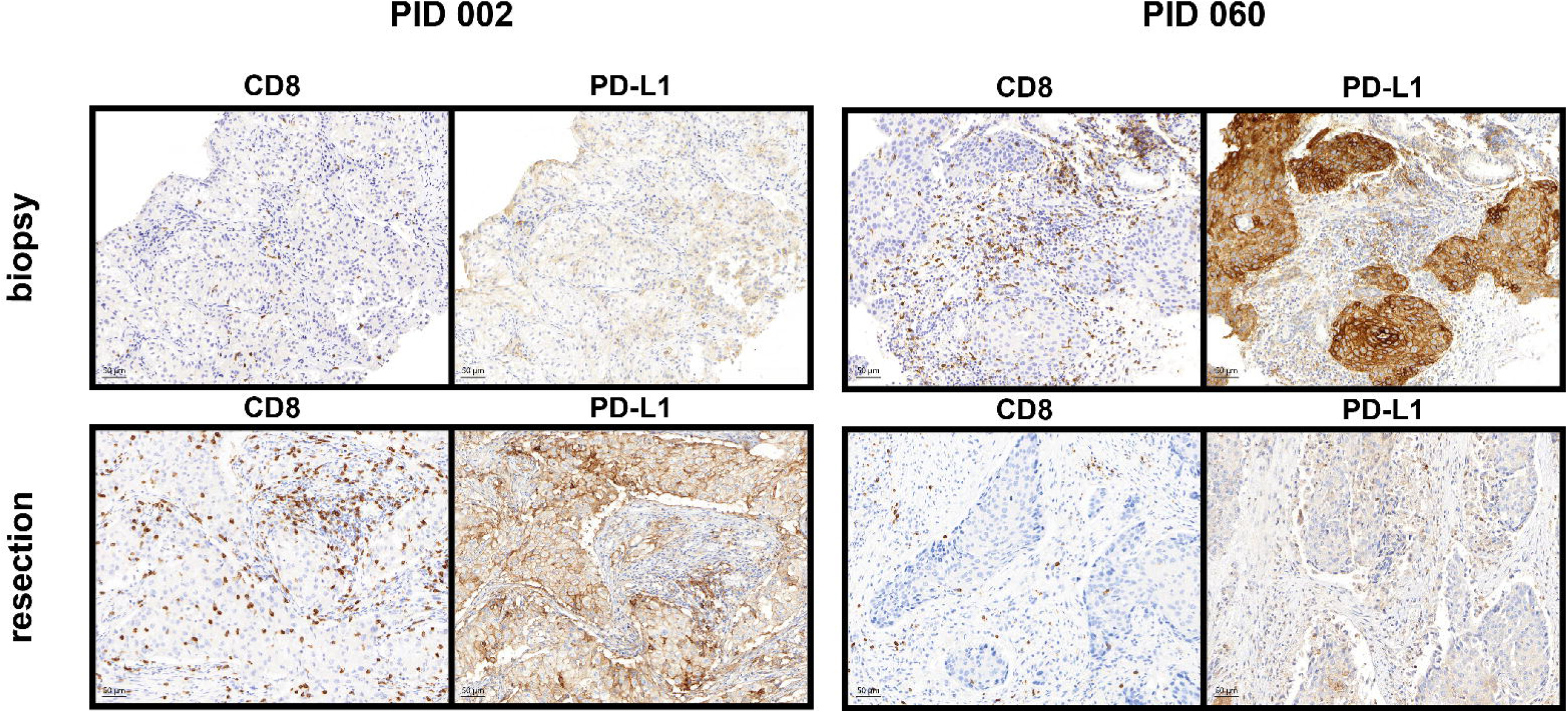
Immunohistochemical slides of two cases in the study cohort with altered PD-L1 expression and TILs density after neoadjuvant therapy. Pre-neoadjuvant and post-neoadjuvant sections stained for CD8 and PD-L1 are represented. Patient 002 changed from 40% TPS to 100% TPS and CD8+ TILs density doubled in the resection specimen. Patient 060 changed from 100% TPS to <1% TPS and CD8+ TILs density halved in the resection specimen. Objective magnification 20x, scalebar 50 µm.

### Higher CD8 TILs density is associated with higher PD-L1 expression

After neoadjuvant therapy, mean CD8 TILs density within the tumor region was 242.45 (IQR 73.11 – 290.32) cells/mm^2^. In the cohort of primary resected cases, mean CD8 TILs density was 252.1 (IQR 98.37 – 314.73) cells/mm^2^. CD8 TILs density was similar between the neoadjuvant and primary resected cohort. After neoadjuvant therapy, a median of 10 (IQR 4 – 21.5) TLS were counted on the selected whole slides. In the primary resected cohort, a median of 9 (IQR 4.5 – 19) TLS were counted. After subgrouping according to histology, the median number of TLS was comparable between histological tumor types and groups. However, there were only 4 cases with active TLS in the neoadjuvant cohort compared to 13 cases in the primary resected control cohort. A higher number of active TLS comparing the cohorts was observed regardless of histological tumor type.

A higher CD8 TILs count was statistically significantly associated with LUAD histology (p_neoadjuvant_ = 0.001, p_control_ = 0.017) and a higher PD-L1 expression (p_neoadjuvant_ = 0.027 R_S_ = 0.23, p_control_ = 0.003 R_S_ = 0.28, Figure 2A). However, when subgrouping according to histological tumor type, PD-L1 expression did no longer significantly correlate with CD8 TILs density in LUAD after neoadjuvant treatment. Similar results were observed when applying the clinical cut-offs at 1% or 50% PD-L1 expression (Figure 2B-C). Among the primary resected cases, PD-L1 positive cases showed a higher CD8 TILs density. After neoadjuvant therapy, this remained true only for non-LUAD tumors. Strong PD-L1 expression correlated with CD8 TILs density in LUAD or non-LUAD tumors in both cohorts. In primary resected cases, tumor size inversely correlated with CD8 TILs density (R_s_ = -0.24, p = 0.011), whereas in cases after neoadjuvant treatment, higher numbers of TLS correlated with higher CD8 TILs density (R_s_ = 0.27, p = 0.009).

**Figure 2:**
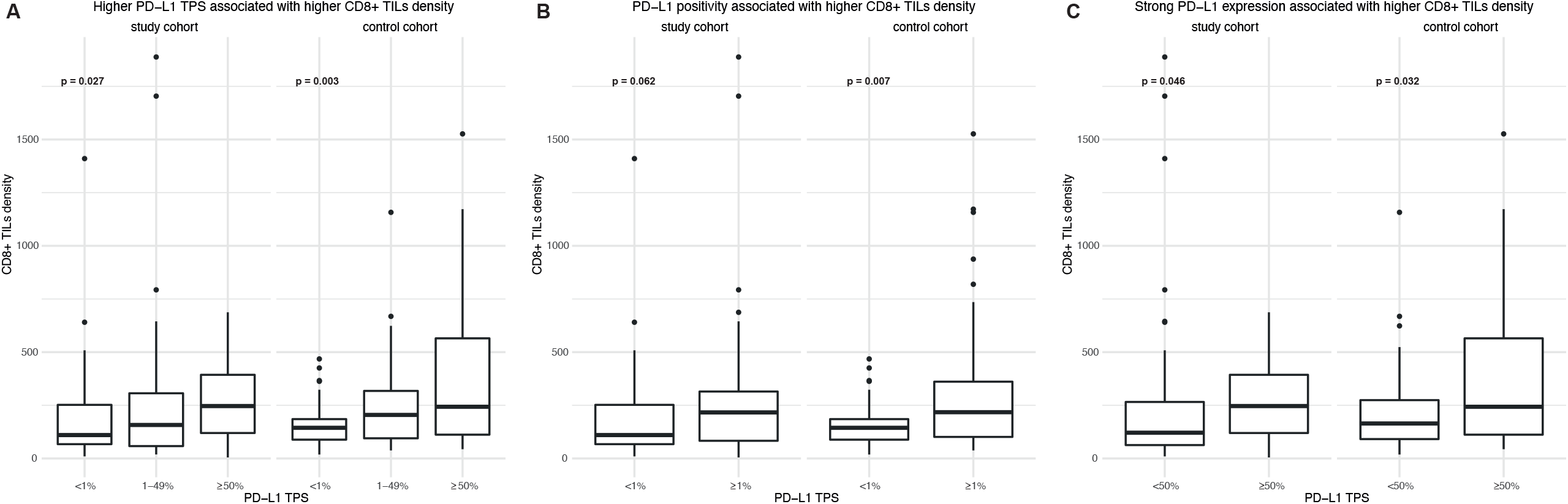
CD8+ TILs density according to PD-L1 expression using the (A) threefold classification, (B) 1% TPS cutoff and (C) 50% TPS cutoff.

CD8 TILs density was significantly lower before neoadjuvant therapy comparing paired samples (p = 0.031, Figure 1, Supplementary figure 4). We performed subgroup analyses to check whether changes of CD8 TILs density were associated with changes in PD-L1 expression. However, changes of CD8 TILs density were only significant in the subgroups of cases with no change of PD-L1 expression regarding the three-fold classification of cut-offs (Supplementary figure 5), presumably due to insufficient sample size in the other subgroups. Furthermore, higher CD8 density before or after neoadjuvant therapy was not associated with an increase of PD-L1 expression.

### Prognostic significance of immune related biomarkers

In the entire study population, PD-L1 expression assessed in resection specimens had no prognostic significance, neither for OS nor for DFS, neither using 1% nor 50% as cut-offs (Supplementary figure 6). In subgroup analyses including only cases after neoadjuvant therapy or primary resected cases, PD-L1 positivity was a prognostic marker for longer OS in the cohort of primary resected NSCLC (p = 0.029, HR 0.5255, 95% CI 0.2924 – 0.9444, Figure 3). Regarding PD-L1 as a dynamic marker, patients with increased PD-L1 expression seemed to have worse 5-year OS, however, not statistically significant (Figure 4).

**Figure 3:**
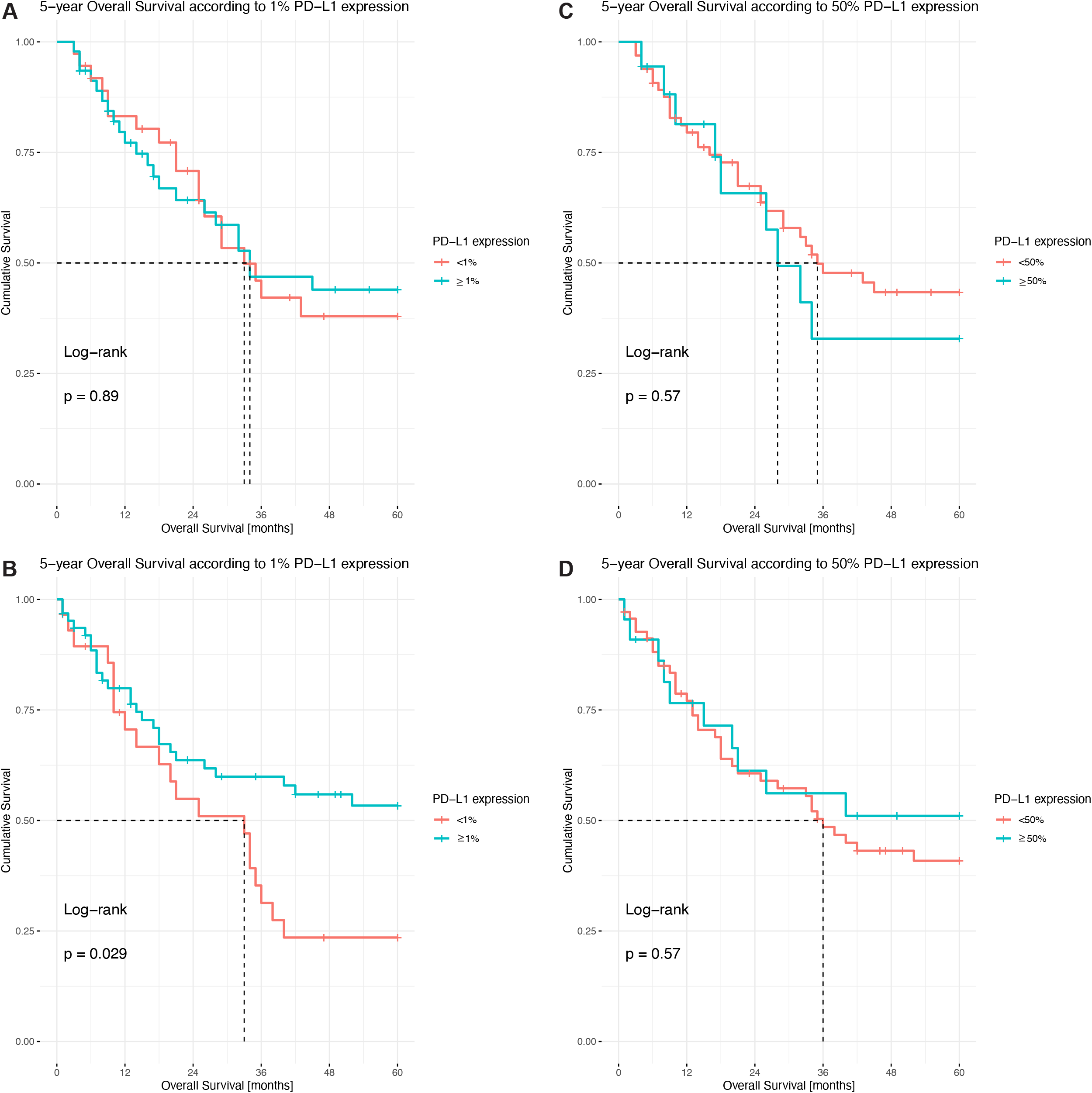
Kaplan-Meier plots of PD-L1 expression as a prognostic biomarker in the (A, C) study cohort and (B, D) control cohort according to the (A, B) 1% TPS cutoff and (C, D) 50% TPS cutoff.

**Figure 4:**
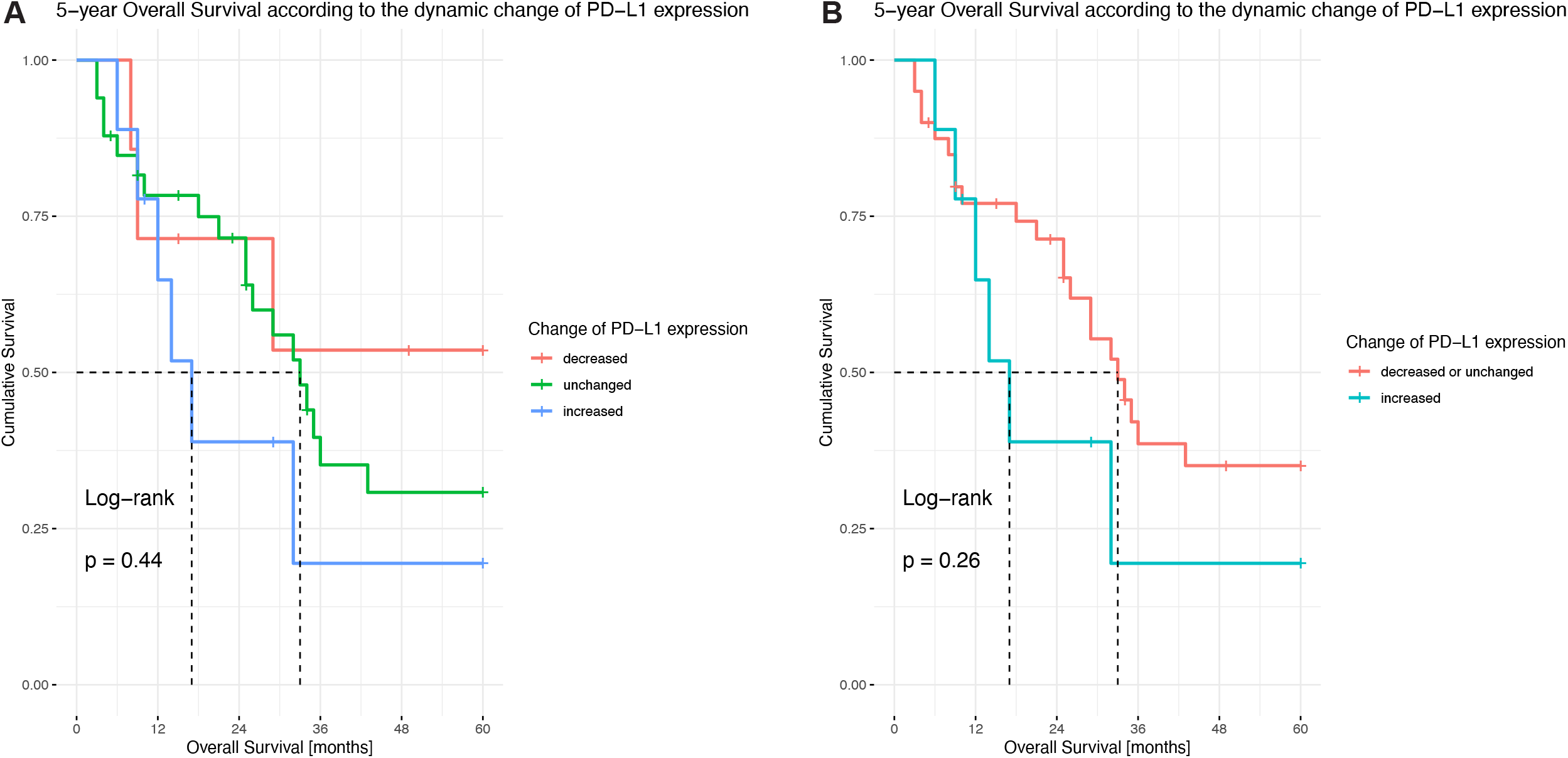
Prognostic importance of the dynamic of PD-L1 change based on the three-fold classification when considering all (A) possibilities or differentiating only between increase and others (B).

We used maximally selected rank statistic to determine the adequate cutoff for CD8 TILs density at 283.18 cells/mm^2^ in the entire study population. Overall, higher CD8 TILs numbers were associated with longer OS (p = 0.014, HR 0.5373, 95% CI 0.3251 – 0.888, Figure 5A) and longer DFS (p = 0.008, HR 0.5707, 95% CI 0.3762 – 0.8656, Figure 5D). In the subgroup analyses, however, it was a positive prognostic factor for OS only in the cohort of neoadjuvant cases (p = 0.029, HR 0.4332, 95% CI 0.1997 – 0.9397, Figure 5B) and for DFS only in the cohort of primary resected cases (p = 0.048, HR 0.5513, 95% CI 0.3043 – 0.9986, Figure 5F).

**Figure 5:**
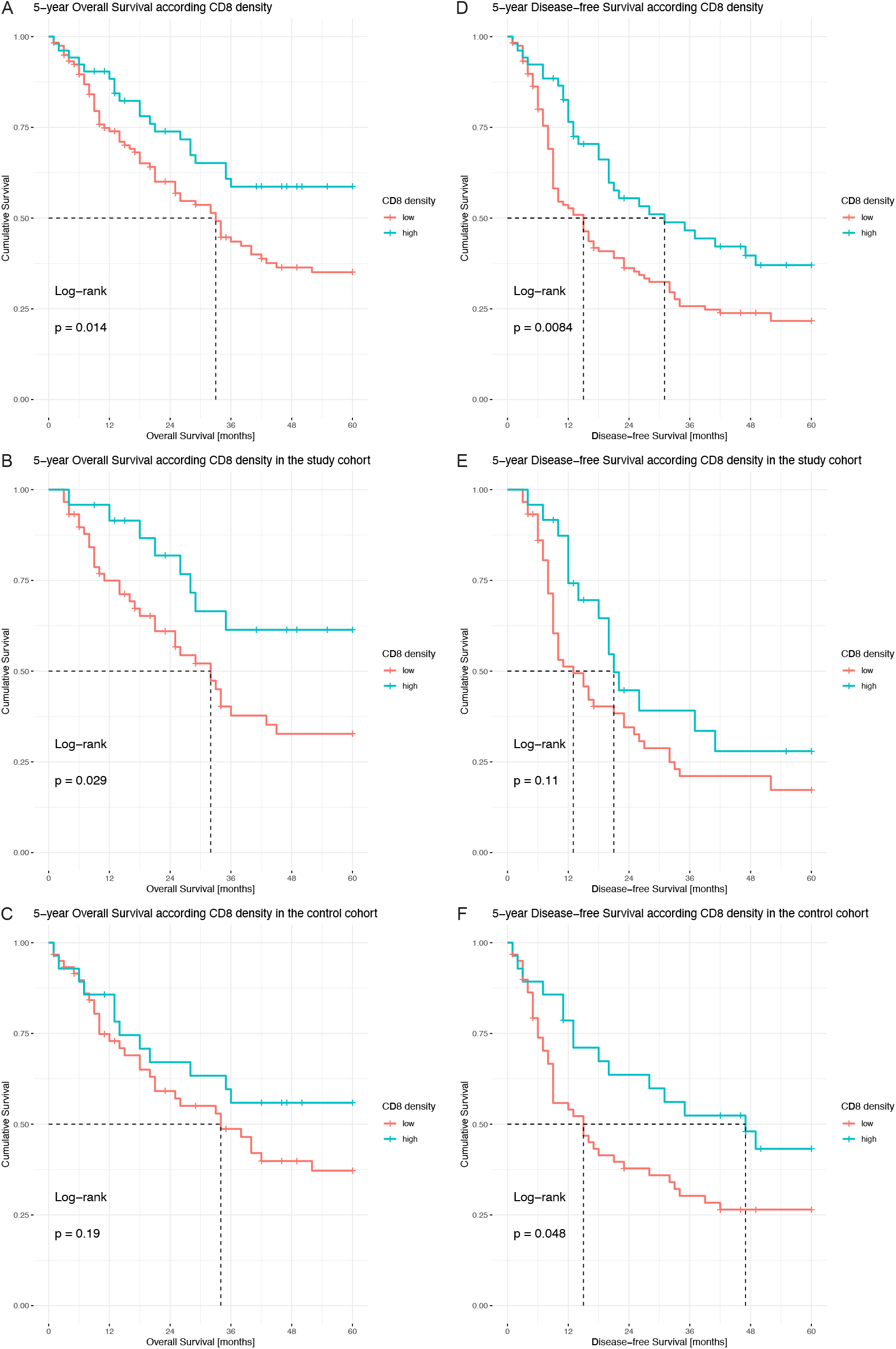
Kaplan-Meier plots of CD8 TILs density as a prognostic biomarker regarding (A-C) OS and (D-F) DFS in the (A, D) entire study population, (B, E) the study cohort and (C, F) the control cohort.

The number of TLS was considered as a continuous variable and it was a prognostic marker considering the entire study population (p = 0.045, HR 0.9833, 95% CI 0.9673 – 0.9996) but not in the sub cohorts (p_neoadjuvant_ = 0.15, p_control_ = 0.15). We investigated the validity of the prognostic significance only in the overall population to achieve sufficient sample size and number of events. As the three immune markers PD-L1 expression, CD8 TILs and TLS correlated significantly, we performed a cox regression analysis for each marker. PD-L1 expression was included using the TPS 1% cut-off only. None of the immune markers was prognostic in the multivariable model for OS (p_PD-L1_ = 0.241, p_CD8_ = 0.368, p_TLS_ = 0.246) or DFS (p_PD-L1_ = 0.054, p_CD8_ = 0.098, p_TLS_ = 0.407) but age and pT4 were consistent prognostic factors in the multivariable models (Supplementary figure 7).

## Discussion

In this study on immune related biomarkers in locally advanced resectable NSCLC, CD8 TILs and TLS were prognostic factors but did not yield additional information to age and TNM in multivariable analyses. CD8 TILs density correlated with PD-L1 expression. PD-L1 expression was not consistently upregulated after neoadjuvant chemotherapy, in line with some, but not all previous studies.(13, 16, 19-22) The effect of chemotherapy on PD-L1 expression in the resectable setting has been previously investigated as summarized in Table 1 (11-22). Most studies reported an increased PD-L1 expression after chemotherapy when assessing paired samples. The importance of the PI3K/ALK pathway in PD-L1 upregulation after chemotherapy was demonstrated in *in vitro* and *in vivo* functional assays (14, 19). Conversely, some studies showed no change or even a decreased PD-L1 expression following chemotherapy (11, 12, 15, 17, 18). In fact, when reassessing the positive studies and considering the absolute number of cases per category (*decrease, no change, increase*), most cases did not change. Of the four studies conducted using an FDA approved antibody, only one concluded an increased PD-L1 expression post-chemotherapy (15, 17, 18, 20). Thus, our study results are in line with previous reports for FDA approved antibodies and suggest that dynamic changes in PD-L1 expression due to chemotherapy are observed only in a minority of tumors. In the majority of our cases (38/57) PD-L1 expression was stable and most of them were negative (23/38 cases) in both the biopsy/cytology specimens and the resection after neoadjuvant therapy. Considering these and previously reported results, it seems unlikely that neoadjuvant chemotherapy induces PD-L1 expression.

Chemotherapy is deemed to improve immunosurveillance by different effects (antigenicity, immunogenicity, susceptibility) (34). This would suggest an upregulation of PD-L1 in tumor cells to evade a strong antitumor response. A potential explanation for the lack of consistent upregulation in clinical samples could be the influence of chemotherapy on the immune microenvironment. Two larger studies have investigated such effects in detail by applying multiplex immunofluorescence and a multiomics approach (16, 35). Both studies describe an increase of immune cells in specimens after neoadjuvant therapy and point out that different subgroups of immune cells are increased depending on the underlying histological tumor type (16, 35). However, the multiomics approach indicated no increase of T-cell receptor richness and clonality, thus failed to suggest increased antigenicity or susceptibility (35). Importantly, these studies did not include paired samples and, especially the study using multiomics, included only few samples in their analyses (n = 10 - 72 depending on experiment) (35).

In order to account for the immune microenvironment, we assessed the CD8 TILs density and number and state of TLS. By applying a semiautomatic approach, we were able to count CD8 TILs in the entire tumor region of the slides used for PD-L1 assessment, in contrast to the published studies usually performing hot spot analysis. Although we confirmed published results indicating an increase of TILs in tumors resected after neoadjuvant therapy, we cannot conclude that chemotherapy increases CD8 TILs densities due to (a) no significant difference in CD8 density between cases resected after neoadjuvant therapy and primary resected cases, (b) biopsies covering much smaller tissue areas than whole slides (thus being more prone to sampling error due to heterogeneity), and (c) higher variance in post-neoadjuvant therapy specimens leading to higher median ranks and means due to outliers (implications for statistical testing).

Another explanation, comparing preclinical and clinical studies, could be the heterogeneity of applied chemotherapeutic regimens, inherent in our real-life cohort approach. Although, most cytotoxic agents have immunosurveillance enhancing effects, these differ considerably (34). Thus, while oxaliplatin and gemcitabine have been shown to promote immunogenic cell death especially via exposure of calreticulin, others do not without addition of radiotherapy (e.g. cisplatin) (34, 36). Furthermore, although Zhang et al. and Fournel et al. suggest an upregulation of PD-L1 via the PI3K/ALK pathway, earlier studies support rather the downregulation of suppressive checkpoints via the STAT pathway (14, 19, 36). In our study, the majority of patients (n = 54) received cisplatin and docetaxel, but only 12/54 received additional radiotherapy (36). On the other hand, the few patients who had received gemcitabine and had available paired samples (n = 2) showed PD-L1-upregulation despite the lack of neoadjuvant radiotherapy, and only 3/11 cases were PD-L1 negative in the resection specimens.

The prognostic power of PD-L1 expression as a double-edged sword has already been described exhaustively in advanced NSCLC and a high PD-L1 expression does not seem to be consistently associated with worse survival (37). Likewise, most studies including tumors after neoadjuvant therapy reported no prognostic importance of PD-L1 expression (11, 15, 18, 20-22). In this study, static PD-L1 expression, evaluated only in the resection specimens, was not prognostic. However, when looking at the dynamic changes, increased PD-L1 expression seemed to confer worse survival, in accordance with previous studies (12, 14, 15, 21). This effect can be explained by PD-L1 expression potentially conveying chemoresistance and promoting proliferation and epithelial to mesenchymal transition (14, 38). High CD8 TILs were always associated with improved OS. This seems to be true even when applying different cut-offs, as most of the published studies used the median, thus a cohort specific cut-off (11, 15, 18, 22, 39). Furthermore, the prognostic impact of a higher CD8 TILs density is a possible explanation for the prognostic benefit of PD-L1 positivity in primary LUSC, due to the positive correlation of PD-L1 expression and CD8 TILs. In our cohort, however, CD8 TILs lost its prognostic relevance in multivariable analyses including age and the pT denominator of the TNM classification. Thus, we cannot confirm the importance of the immune pattern as a complementary factor for survival prediction, as proposed by Remark and colleagues (11).

This retrospective study assessed the impact of chemotherapy on biomarkers for ICB in a real-life cohort resected after neoadjuvant therapy over a period of 16 years. The availability of paired samples for 57 patients is comparable to prior studies but the addition of a matching cohort of primary resected locally advanced NSCLC allowed the validation of identified effects. In contrast to most published studies, we evaluated PD-L1 expression according to the current diagnostic recommendations and using an FDA-approved antibody assay for companion diagnostics (40-42). Furthermore, we included the impact on the immune microenvironment by evaluating CD8 TILs and TLS. By applying new techniques enabled by digital pathology approaches, we could reliably assess CD8 TILs in the same area as the PD-L1 expression and were not restricted to hotspot analyses.

Nevertheless, our study has limitations inherent to its retrospective and “real-life” character. Especially compared to clinical studies investigating the effect of radio-chemotherapy, patients with different chemotherapeutic regimens were included (15, 18). Although patients with changed PD-L1 expression after neoadjuvant therapy did not differ regarding duration of neoadjuvant therapy or therapy free interval between the last cycle of neoadjuvant therapy and resection, especially these differences in duration of therapy and therapy free interval need to be accounted for when interpreting our results.

In conclusion, our results support the hypothesis of dynamic effects of neoadjuvant chemotherapy on PD-L1 expression and CD8 TILs. Overall, the majority of our cases were PD-L1 negative and showed no increase in PD-L1 expression after neodjuvant therapy. Nevertheless, in those cases with increased PD-L1 expression after neoadjuvant therapy, 5-year OS was shorter compared to patients with no change or decreased PD-L1 expression. This result could be visually appreciated, but lacked statistical significance, most probably due to the very low number of cases. Subsequent clinical trials are warranted in order to determine if PD-L1 retesting should be performed after neoadjuvant therapy due to therapeutic implications of an altered PD-L1 expression.

## Supporting information

REMARK checklist

All supplementary data to this paper

## Data Availability

Anonymized detailed clinico-pathological data and the R-Script used for data analysis are available upon request to the authors.

## Acknowledgment

The authors are very thankful for the expert technical support of the Translational Research Unit of the Institute of Pathology, University of Bern. We acknowledge the Cancer registry Bern for acquiring extensive follow-up data of the patients.

## Additional Information

### Authors’ contribution

Philipp Zens: Conceptualization, Data curation, Formal analyses, Funding acquisition, Methodology, Visualization, Writing – original draft, Writing – Review & Editing; Corina Bello: Data Curation, Writing – Review & Editing; Amina Scherz: Data Curation, Writing – Review & Editing; Michael von Gunten: Resources, Writing – Review & Editing; Adrian Ochsenbein: Resources, Writing – Review & Editing; Ralph A. Schmid: Resources, Writing – Review & Editing; Christina Neppl: Resources, Writing – Review & Editing; Sabina Berezowska: Conceptualization, Data curation, Formal analyses, Funding acquisition, Methodology, Project administration, Resources, Supervision, Writing – original draft, Writing – Review & Editing

### Ethics approval and consent to participate

This study was approved by the Cantonal Ethics Committee of the canton of Bern waiving the requirement of informed consent (KEK 2017-00830). The study was performed in accordance with the Declaration of Helsinki.

