## Supplementary material for "The effect of neoadjuvant therapy on PD-L1 expression and CD8 lymphocyte density in non-small cell lung cancer": All supplementary data to this paper

| **Element** | **Title** | **Page** |
| --- | --- | --- |
| Figure 1 | Overall and disease-free survival of the study population | 2 |
| Figure 2 | Time-dependent change of marker expression | 3 |
| Figure 3 | Semi-automated lymphocyte detection | 4 |
| Figure 4 | Per patient comparison of preneoadjuvant and postneoadjuvant CD8+ TILs density | 5 |
| Figure 5 | CD8 TILs according to PD-L1 change (based on three tier classification of PD-L1) | 6 |
| Figure 6 | Prognostic significance of PD-L1 in locally advanced resectable NSCLC | 7 |
| Figure 7 | Multivariable cox proportional hazard models for prognostic markers | 9 |
| Table 1 | Comparison of manual and automatic TILs detection | 10 |
| Table 2 | PD-L1 expression according to smoking status | 11 |
| Table 3 | Cases with changed PD-L1 expression after neoadjuvant therapy | 12 |
| Table 4 | Association of change of PD-L1 with clinico-pathological parameters | 13 |

### Supplementary figure 1: Overall and disease-free survival of the study population

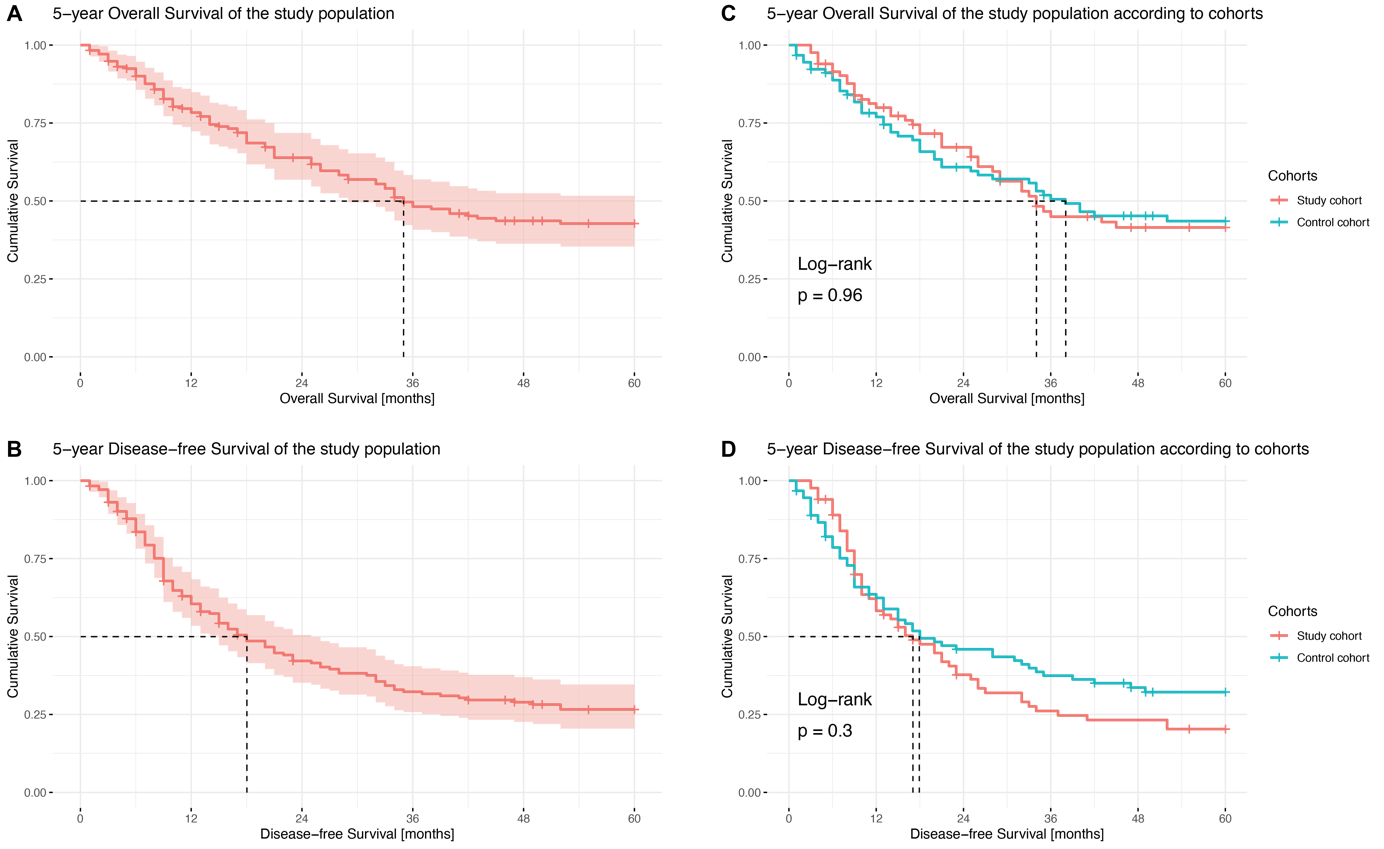

Figure 1: (A) Overall survival and (B) disease-free survival of the study population and (C, D) according to the corresponding sub cohort.

### Supplementary figure 2: Time-dependent change of marker expression

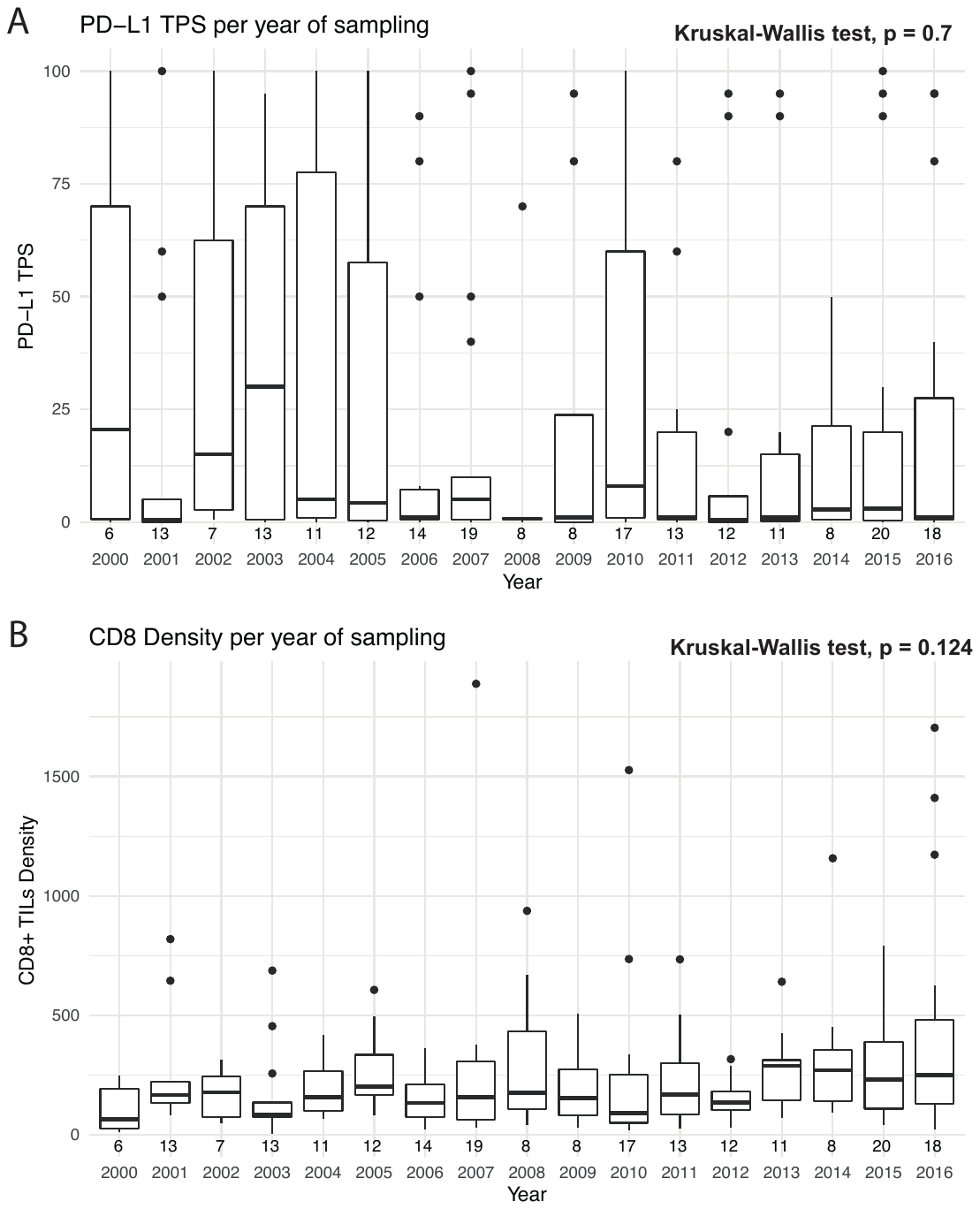

Figure 2: Distribution of (A) PD-L1 and (B) CD8 expression per year during the period of observation. Overall, there is no difference in distribution indicated by p > 0.05 using the Kruskal-Wallis test. The number of observations per year is indicated below each boxplot.

### Supplementary figure 3: Semi-automated lymphocyte detection

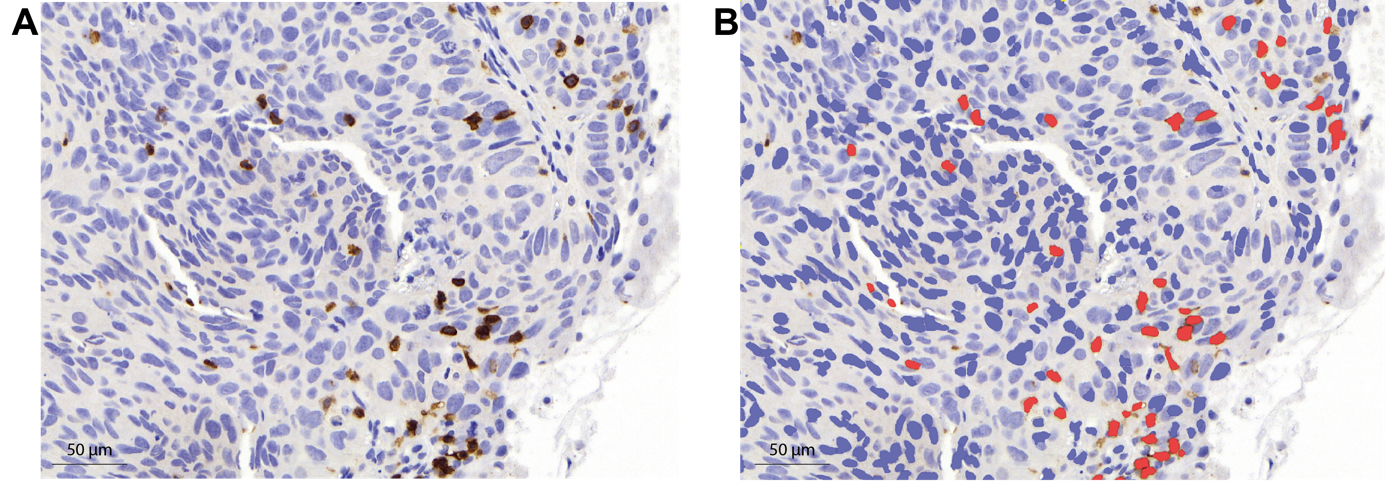

Figure 4: Lung tumor tissue sections immunohistochemically stained for CD8 were analyzed using QuPath. (A) The original image shows scattered lymphocytes stained in brown. (B) The same section with an overlay of the automatically detected and classified cells, CD8+ lymphocytes are marked in red.

### Supplementary figure 4: Per patient comparison of preneoadjuvant and postneoadjuvant CD8+ TILs density

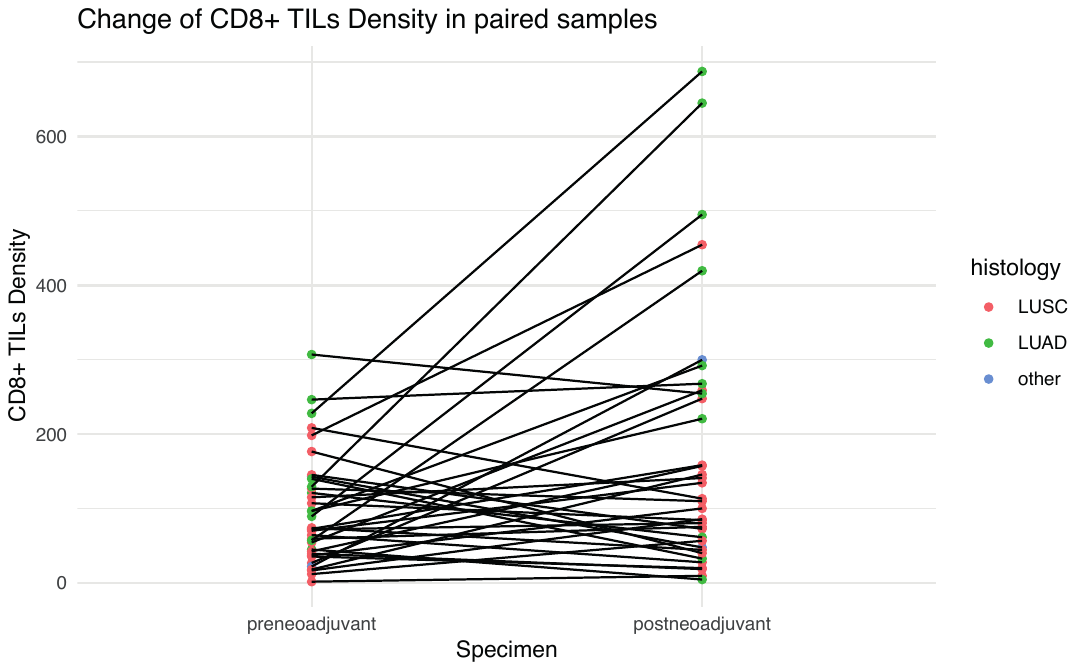

Figure 3: Change of individual CD8+ TILs density in paired specimen. The histologies indicated by color indicate a potential bigger positive change in lung adenocarcinomas compared to non-adenocarcinomas. LUSC: lung squamous cell carcinoma, LUAD: lung adenocarcinoma.

### Supplementary figure 5: CD8 TILs according to PD-L1 change (based on three tier classification of PD-L1)

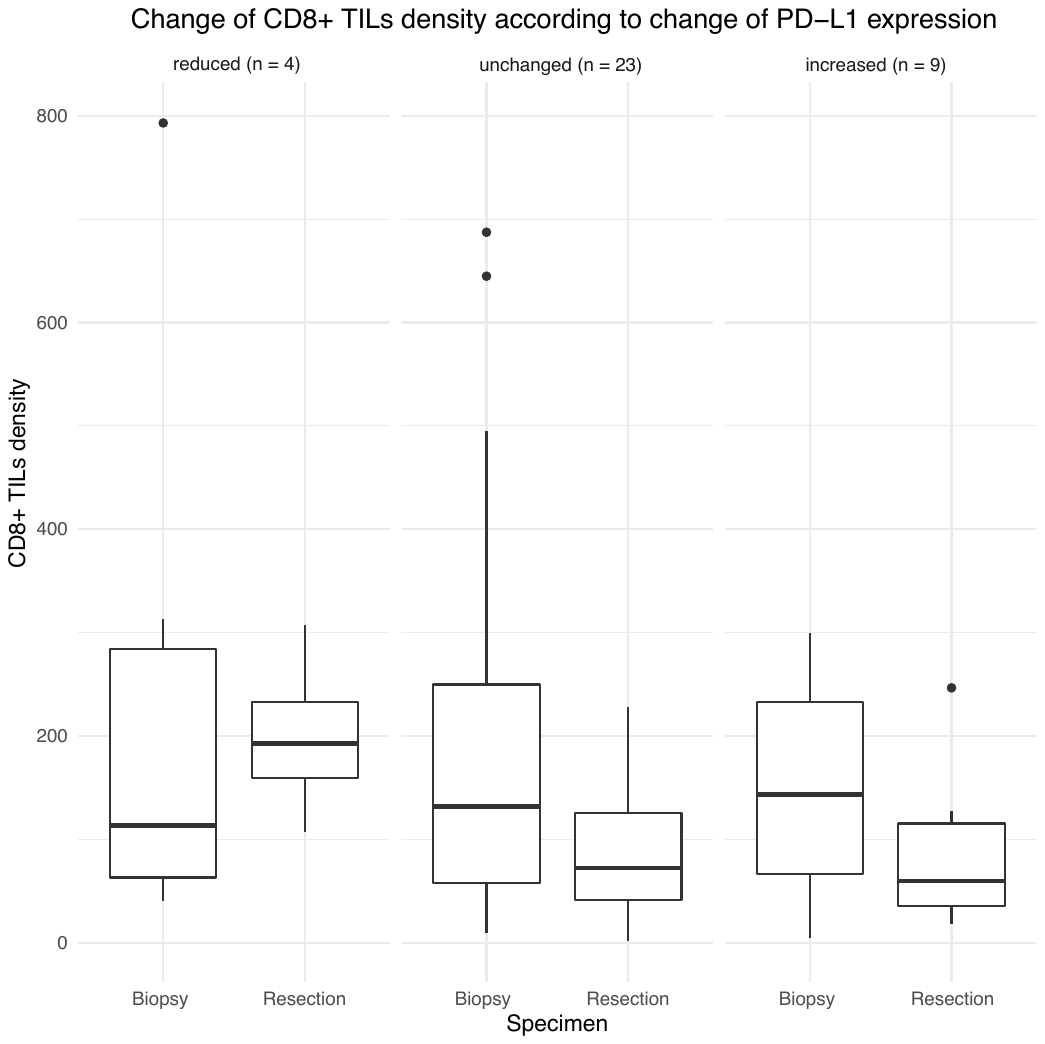

*Figure 5: Change of CD8+ TILs density according to change of PD-L1 TPS in paired specimens. Significant decrease of CD8+ TILs (p = 0.012) was detected only in the group of unchanged PD-L1 expression, probably due to sufficient sample size.*

### Supplementary figure 6: Prognostic significance of PD-L1 in locally advanced resectable NSCLC

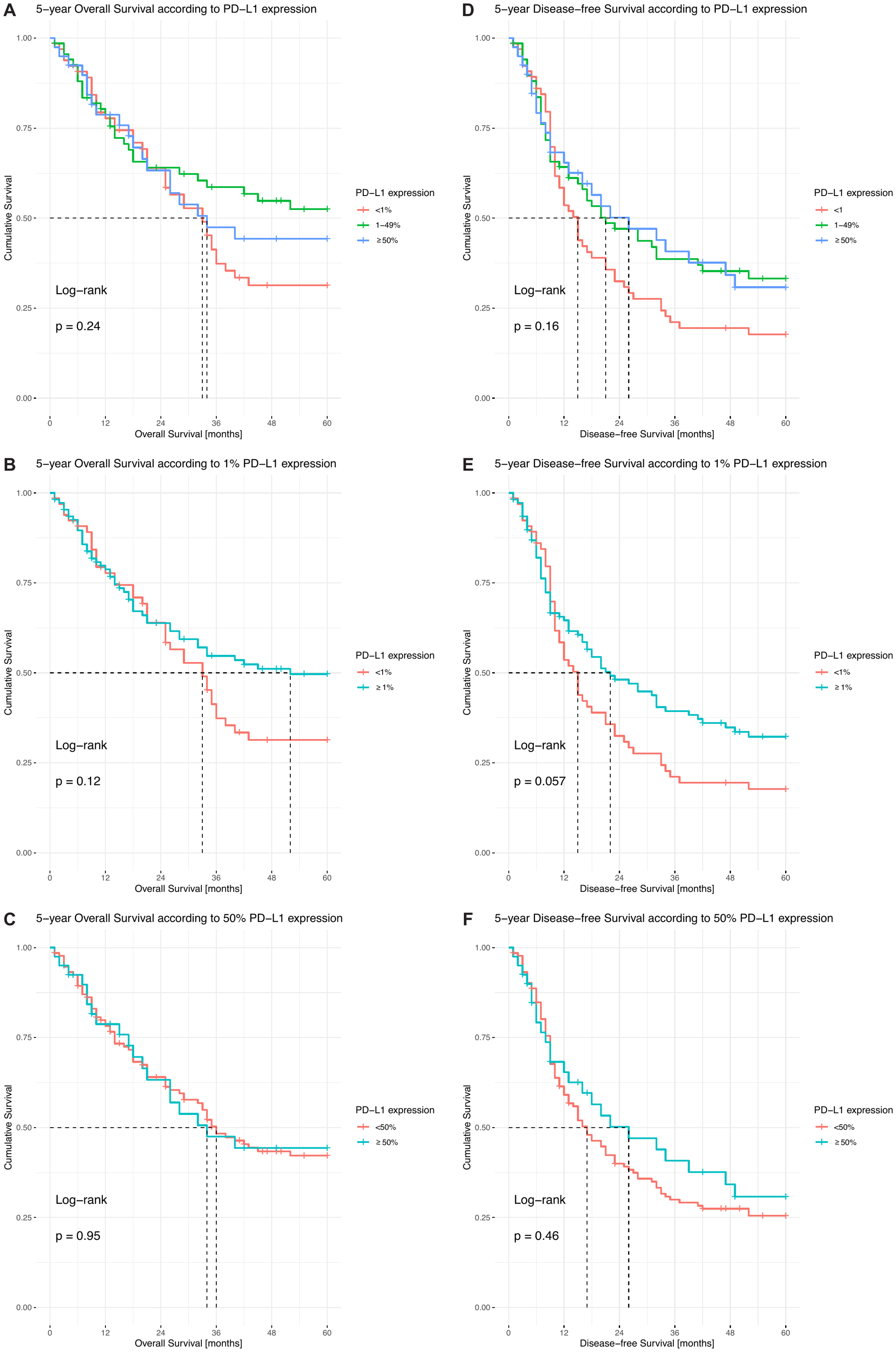

*Figure 6: Kaplan Meier curves of (A, B, C) OS and (D, E, F) DFS according to PD-L1 TPS (A, D) threefold, (B, E) 1% cut-off or (C, F) 50% cut-off.*

### Supplementary figure 7: Multivariable cox proportional hazard models for prognostic markers

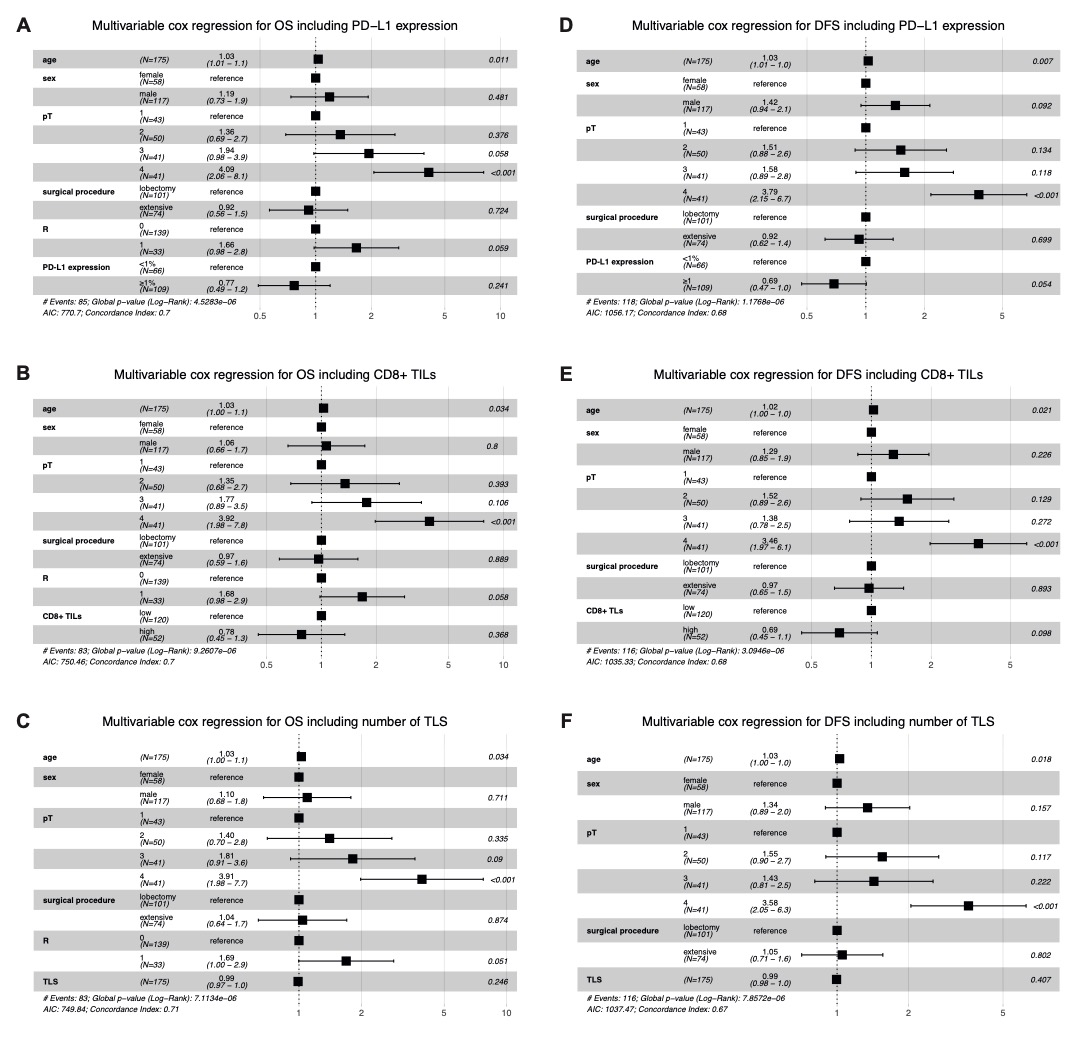

Figure 7: Forest plots integrating immune biomarkers in a multivariable cox proportional hazard model for (A-C) overall survival and (D-F) disease-free survival including (A, D) PD-L1 expression, (B, E) CD8+ TILs and (C, F) the number of tertiary lymphoid structures.

### Supplementary table 1: Comparison of manual and automatic TILs detection

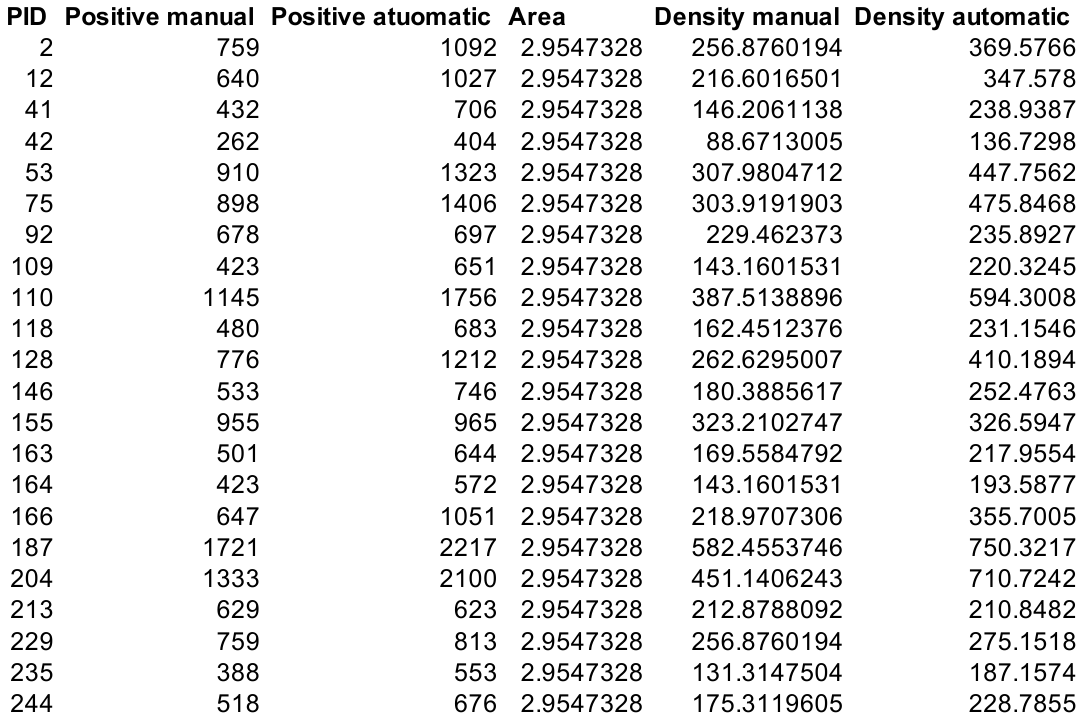

Table 1: Comparison of manually and automatically counted infiltrating CD8+ lymphocytes.

### Supplementary table 2: PD-L1 expression according to smoking status

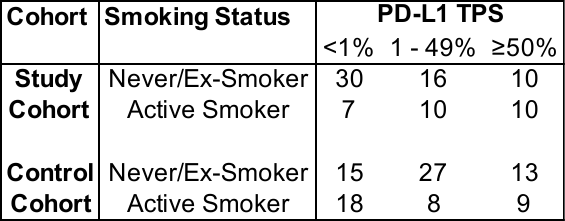

*Table 2: Association of PD-L1 and smoking status.*

### Supplementary table 3: Cases with changed PD-L1 expression after neoadjuvant therapy

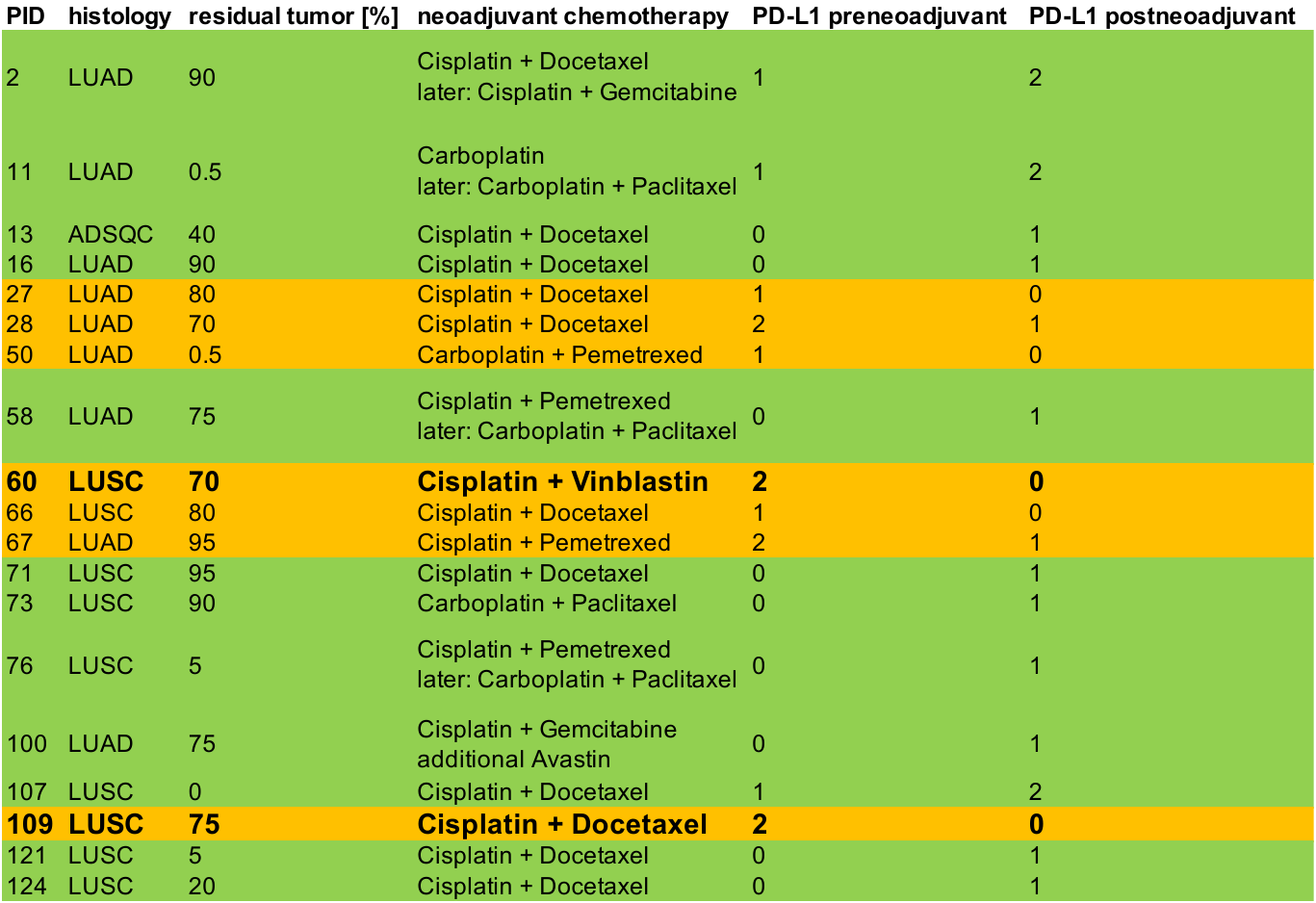

Table 3: Patients with different PD-L1 expression after neoadjuvant therapy. Green cases showed higher and orange cases lower PD-L1 expression after neoadjuvant therapy. Two cases (bold, larger font) showed a drastic decrease of PD-L1 expression from >95% (PID60) and 70% (PD109) to <1% respectively.

### Supplementary table 4: Association of change of PD-L1 with clinico-pathological parameters

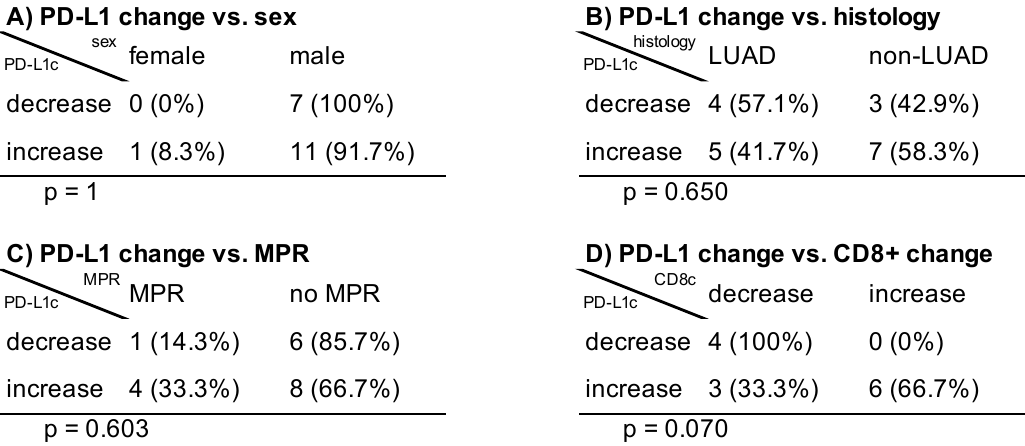

Table 4: Assessment of potential associations of (A) sex, (B) histology, (C) presence or absence of major pathological response (MPR) and (D) change in CD8+ TILs density with change in PD-L1 TPS. PD-L1c: change of PD-L1 TPS (cut-offs 1% or 50%), LUAD: lung adenocarcinoma, CD8c: change of CD8+ TILs density. P-values are reported using Fisher’s exact test.
